# Intraoperative adjustment of radiographic standard projections of the spine: Interrater- and intrarater variance and consequences of ‘fluoro-hunting’ considering time and radiation exposure – A cadaveric study

**DOI:** 10.1101/2022.02.12.22270884

**Authors:** Eric Mandelka, Jan El Barbari, Lisa Kausch, Maxim Privalov, Paul Alfred Grützner, Sven Y. Vetter, Jochen Franke

**Author notes:** **Corresponding Author** Eric Mandelka. <u>Declarations</u>. **Ethics Approval** The study was reviewed and approved by the ethics committee of the Rhineland-Palatine Medical Association (application number 2020-15423). All procedures performed were in accordance with the ethical standards of the institutional and/or national research committee and with the 1964 Helsinki Declaration and its later amendments or comparable ethical standards. **Informed Consent** The human specimens were provided by Rimasys GmbH (Cologne, Germany). The body donors consented to the donation of their bodies for research purposes during their lifetime. **Availability of Data and Material** All data and statistics are available on reasonable request from the corresponding author. **Authors’ Contribution** Eric Mandelka: Data curation, Formal analysis, Writing - original draft, Writing – review and editing. Jan El Barbari: Investigation, Data curation, Visualization, Writing – review and editing. Lisa Kausch: Data Curation, Formal analysis, Writing – review and editing. Maxim Privalov: Conceptualization, Writing – review and editing. Sven Y. Vetter: Data Curation, Writing – review and editing. Paul A. Grützner: Supervision, Writing – review and editing. Jochen Franke: Methodology, Writing – original draft. Writing – review and editing. All authors read and approved the final manuscript.

## Abstract

**Background:** For the acquisition of intraoperative fluoroscopic images, standard projections have to be manually adjusted. This process resembles a trial-and-error process and is therefore time-consuming and leads to increased radiation exposure for both patient and staff. In addition, the standard projections adjusted are subject to intra- and interindividual variance. However, to date, only very limited data exist in the literature quantifying the time and radiation exposure caused by the process of manually setting standard projections as well as the intra- and interindividual variance for the manual adjustment of standard projections.

**Material and Methods:** A.p. and lateral standard projections of the vertebral bodies of two fresh-frozen specimen were manually adjusted by two examiners with a different level of experience using a mobile C-arm. The time needed for manual adjustment as well as the number of X-ray shots acquired and the radiation dose caused during this process were documented. Intra- and interindividual variance of the central beam, the orbital rotation and angulation of the C-arm was analyzed.

**Results:** The median time needed was 75.9s, with no significant difference between the examiners (p=0.13). 7.1 x-ray images were acquired in average to reach subjective satisfaction with the standard projection with significantly more x-ray shots for the lateral standard (p=0.04) and for the examiner with less experience (p<0.001). Accordingly, the dose caused was more than 50% higher than for the experienced examiner (p=0.01). Mean interindividual variance of the central beam was 7.6° while the intraindividual variance was 4.2°.

**Conclusion:** In summary, this study investigated the interrater and intrarater variance for standard manual level setting in the thoracic and lumbar spine. Additionally, we were able to quantify the time and number of radiographs required for this procedure for different levels of experience, as well as the resulting radiation dose.

## Introduction

When taking fluoroscopic images, the adjustment of standard projections is part of the standard procedure. This is even more important intraoperatively, for example to check fracture reduction and implant position. The manual setting of standard projections with the multiple degrees of freedom available with a mobile C-arm, especially if the surgeon lacks experience, resembles a trial-and-error process. During this process multiple radiographic images are acquired with corrections being made to patient positioning or positioning of the device in between until a correct setting is achieved. In addition to the time involved, this so-called ‘fluoro hunting’ leads to increased radiation exposure for both the patient and the staff[1, 2].

A clinical example is the adjustment of standard projections of single vertebral bodies in the context of fluoroscopic-controlled pedicle screw placement. In this setting, surgeons are exposed to high levels of radiation, even compared to other surgeons[3, 4]. However, to date, only very limited data exist in the literature quantifying the time and radiation exposure caused by the process of manually setting standard projections.

The result, the manually adjusted standard projection, is also highly dependent on the examiner. On the one hand, this is due to non-existent or inconsistent standards and the lack of widespread use of these. On the other hand, however, this variance is the result of the manual setting, especially if it is performed by less experienced operators. To date, no studies have been described in the literature that examine intra- and interrater variance in the setting of standard projections.

Yet, these data are of particular importance with regard to existing efforts to support the surgeon in the manual setting of standard projections with software-technical solutions[5, 6]. For example, Kausch et al. published an approach to automatically suggest the necessary corrections of the C-arm positioning to the surgeon based on a first fluoroscopic image[7]. This approach might only be an interim solution until the setting of the correct standard projections is fully automated based on a first image.

However, for these methods to be transferred to the clinical approach, quality control of the automatically set standard projections must take place first. For this purpose, the variance of the automatically set standard projections needs to be minimized to ensure that the results always meet clinical demands. To quantify these demands, knowledge of variance of manual setting of standard projections is essential.

Therefore, this experimental study will use the example of the spine to investigate the intra- and interrater variance in the manual adjustment of standard radiographic projections. In addition, the radiation exposure caused during this process and the time required will be quantified for examiners with different levels of experience.

## Methods

Two fresh-frozen human specimens consisting of torso with pelvis were positioned in prone position on two radiolucent carbon tables (Fig. 1). Subsequently, the standard radiographic projections antero-posterior (ap) and lateral (lat) for the vertebral bodies Th1 to L5 were manually adjusted with two mobile C-arms (Cios Spin, Siemens Healthineers, Erlangen Germany). This was done by two examiners, an experienced spine surgeon (in the following referred to as examiner 1) and a orthopedics and trauma surgery resident (in the following referred to as examiner 2). The examiners worked simultaneously at two different workstations so that they were blinded to the exact settings of the other examiner. After having finished the adjustment on the first specimen, workstations were swapped, and the examiners adjusted the standard projections for the respective other specimen to analyze interrater variance. Subsequently, to investigate intrarater variance, examiner 1 once again adjusted the standard projections on the first specimen.

**Fig. 1.**
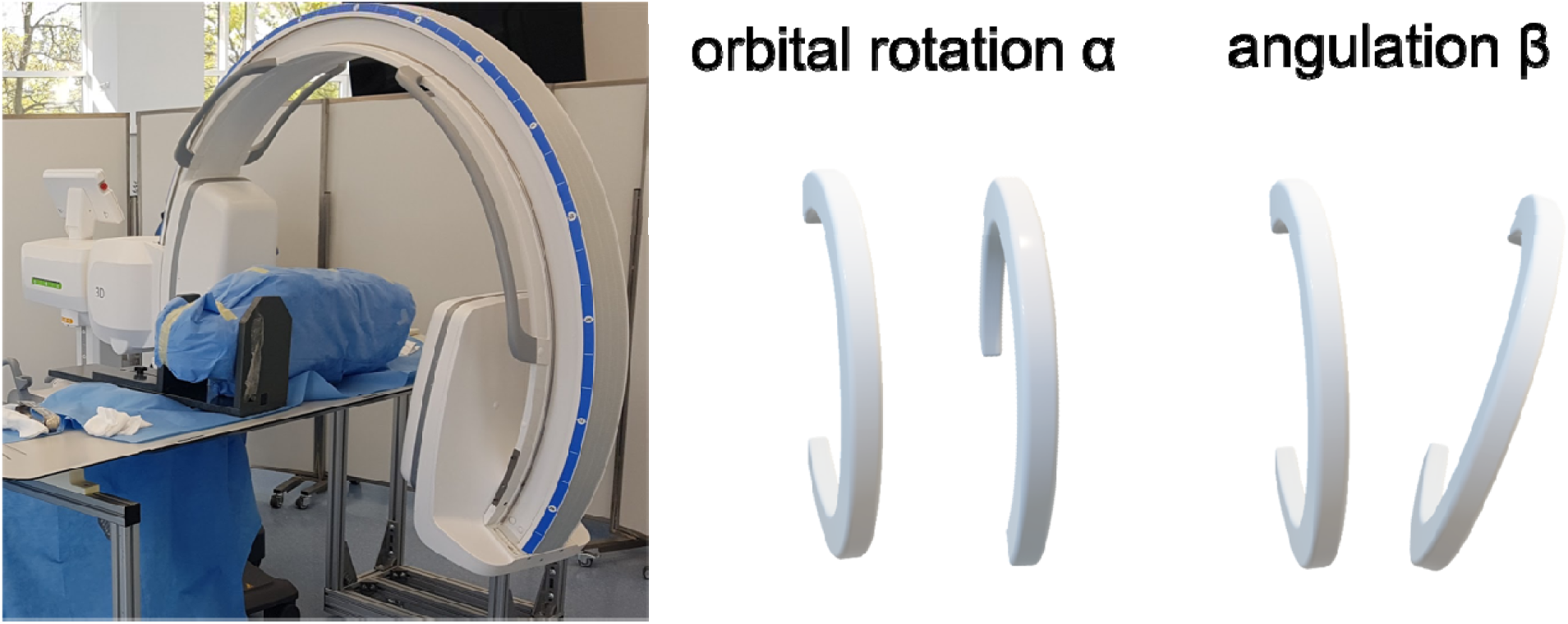
Experimental setup with specimen and mobile C-arm and visualization of the degrees of freedom investigated.

The standard projections were defined as follows:

- a.p.:
  ○ respective vertebra in central beam
  ○ spinous process centralized
  ○ symmetrical visualization of vertebral arches and transverse processes
  ○ superior and inferior endplate struck orthograde (visible as one line)
  ○ good visibility of intervertebral spaces
- lateral:
  ○ respective vertebra in central beam
  ○ spinous process clearly visible
  ○ superior and inferior endplate struck orthograde (visible as one line)
  ○ good visibility of intervertebral spaces

By two other investigators present, the following variables were collected during the experiments:

- The time required to adjust the standard projections, measured from the first change in C-arm positioning from neutral position to the subjectively correct adjustment of the standard projection.
- The number of X-ray single images taken in each case until the standard projection was subjectively set correctly. Radiographs obtained to identify the specific vertebra were not included in the count.
- The radiation dose caused during each individual setting of the standard projections, expressed as a dose area product (in mGy*cm^2^) from the examination protocol of the C-arm.

For the evaluation of intrarater and interrater variance in the correctly set standard projections, the following parameters were documented for the image data acquired:

- dθ: variance of the angle of the central beam (in °)
- dα: variance of the orbital rotation angle (in °)
- dβ: variance of the angulation angle (in °)

Radiation protection measures in terms of lead aprons with thyroid shields as well as mobile lead walls were followed. Dosimeters to record radiation exposure were worn by all participants throughout the duration of the experiments.

The descriptive statistical analysis on the dataset tabulated in Excel (Version 16.57, Microsoft Corporation) was conducted using Prism 8 (Graphpad Software, Inc.).

All data were tested for normal distribution using Kolmogorov-Smirnov test. Descriptive statistics are shown as means and standard deviations (SD) for normally distributed data, and medians and interquartilary range (IQR) for data without a Gaussian distribution. To facilitate comparison with literature results, mean values were also reported in Table 1, although the data were not normally distributed. The Wilcoxon matched-pairs signed-rank test was used to analyze the central tendencies for the time needed, the number of radiographs needed and the radiation dose cause. The significance level was set at p<0.05.

**Tab. 1:** Mean with standard deviation (SD) and median with IQR for time and number of x-rays needed to adjust standard projection and radiation dose caused per vertebra.

|  | Overall | Examiner 1 | Examiner 2 |  |
| --- | --- | --- | --- | --- |
| Time [s] |  |  |  |  |
| overall | 87.3 (SD 66.5) | 79.7 (SD 64.6) | 94.9 (SD 67.9) | p=0.13 |
|  | 75.9 (IQR 63.8) | 72.4 (IQR 55.1) | 81.1 (IQR 76.9) |  |
| a.p. | 91.0 (SD 48.3) | 83.8 (SD 33.4) | 98.2 (SD 59.3) |  |
|  | 78.8 (IQR 49.0) | 76.6 (IQR 38.2) | 81.1 (IQR 54.8) |  |
| lat | 83.6 (SD 80.9) | 75.6 (SD 85.6) | 91.7 (SD 76.4) |  |
|  | 60.1 (IQR 87.9) | 49.2 (IQR 72.9) | 77.1 (IQR 109.3) |  |
| # X-ray single images [n] |  |  |  |  |
| overall | 7.1 (SD 5.4) | 5.5 (SD 4.2) | 8.8 (SD 6.0) | p<0.001 |
|  | 6.0 (IQR 6.0) | 5.0 (IQR 3.8) | 7.0 (IQR 7.0) |  |
| a.p. | 7.7 (SD 4.6) | 5.8 (SD 2.5) | 9.6 (SD 5.3) |  |
|  | 6.0 (IQR 4.0) | 6.0 (IQR 2.3) | 8.0 (IQR 6.3) |  |
| lat | 6.6 (SD 6.2) | 5.2 (SD 5.3) | 8.0 (SD 6.7) |  |
|  | 4.5 (IQR 7.6) | 3.5 (IQR 5.0) | 6.0 (IQR 9.3) |  |

| Dose [$\mu\text{Gy}\cdot\text{cm}^2$] | | | | |
| --- | --- | --- | --- | --- |
| overall | 76.0 (SD 113.5) | 62.6 (SD 114.9) | 89.3 (SD 111.2) | p=0.01 |
|  | 34.5 (IQR 66.3) | 29.7 (IQR 38.5) | 46.5 (IQR 80.9) |  |
| a.p. | 39.7 (SD 31.0) | 29.7 (SD 28.9) | 49.7 (SD 30.3) |  |
|  | 32.1 (IQR 32.4) | 25.0 (IQR 19.9) | 43.0 (IQR 30.1) |  |
| lat | 112.3 (SD 149.3) | 95.5 (SD 154.1) | 129.0 (SD 144.7) |  |
|  | 44.7 (IQR 125.1) | 34.6 (IQR 108.8) | 86.2 (IQR 162.9) |  |

## Results

### Time needed for adjustment of standard projections

The time needed for the adjustment of standard projections ranged from 5.2 to 473.2s with a median of 75.9s. Examiner 1 tended to set the standard projections faster than examiner 2, however, the difference was not statistically significant (p=0.13, Table 1). Both investigators tended to require more time to adjust standard lateral projections than for standard ap projections (p=0.11, Fig. 2).

**Fig. 2.**
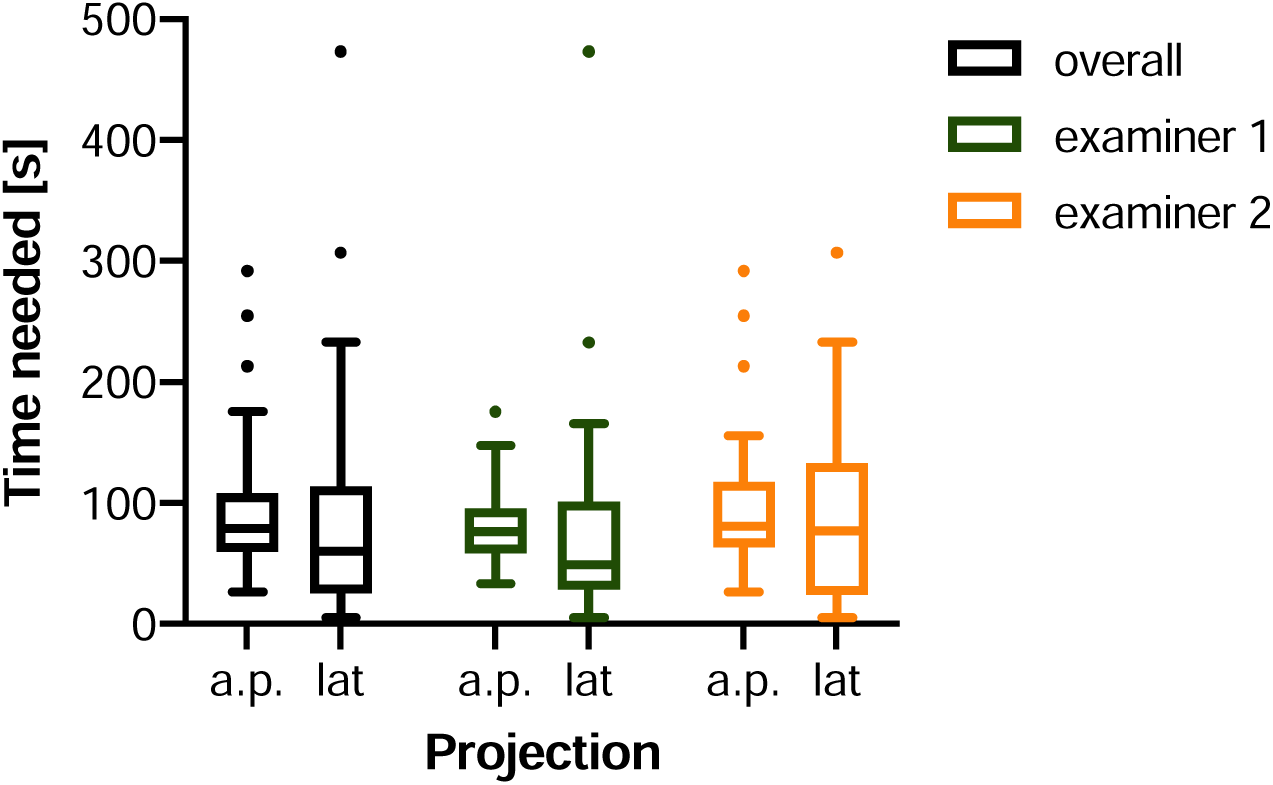
Time needed for the manual adjustment of a standard projection for a single vertebral body.

The number of x-ray shots that was done to reach subjective satisfaction with the standard projection adjusted was significantly higher for ap than for lateral standard (p=0.04). The difference between both examiners reached statistical significance with a median of 5 and 7 shots needed, respectively (p<0.001, Fig. 3).

**Fig. 3.**
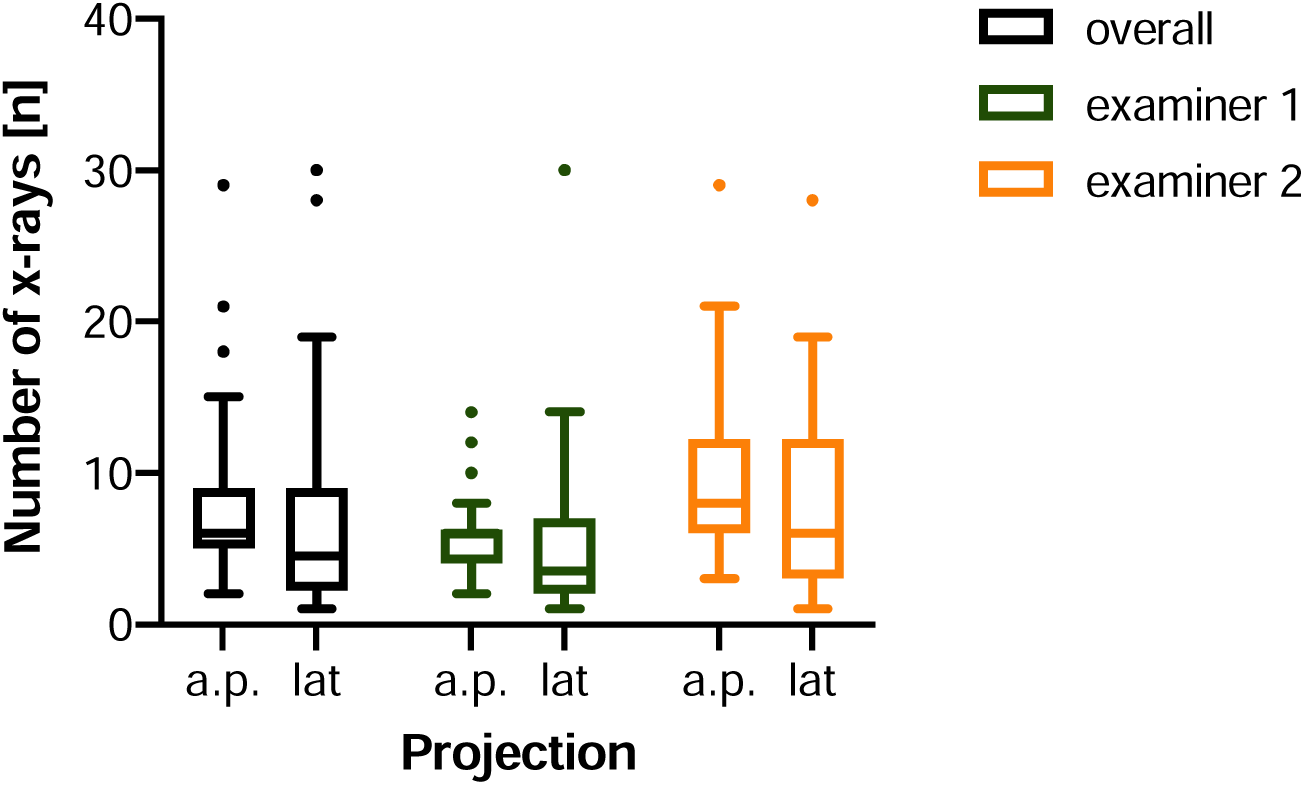
Number of single images taken for the manual adjustment of a standard projection for a single vertebral body.

The median dose per vertebra caused by those shots for both investigators was 34.5 μGy*cm^2^. When comparing the dose caused by the two examiners, the difference showed to be significant with the median of examiner 2 being more than 50% higher than the median of examiner 1 (p=0.01). The dose caused by the adjustment of lateral standard projections was significantly higher than for ap standard projections (p<0.001, Fig. 4).

**Fig. 4.**
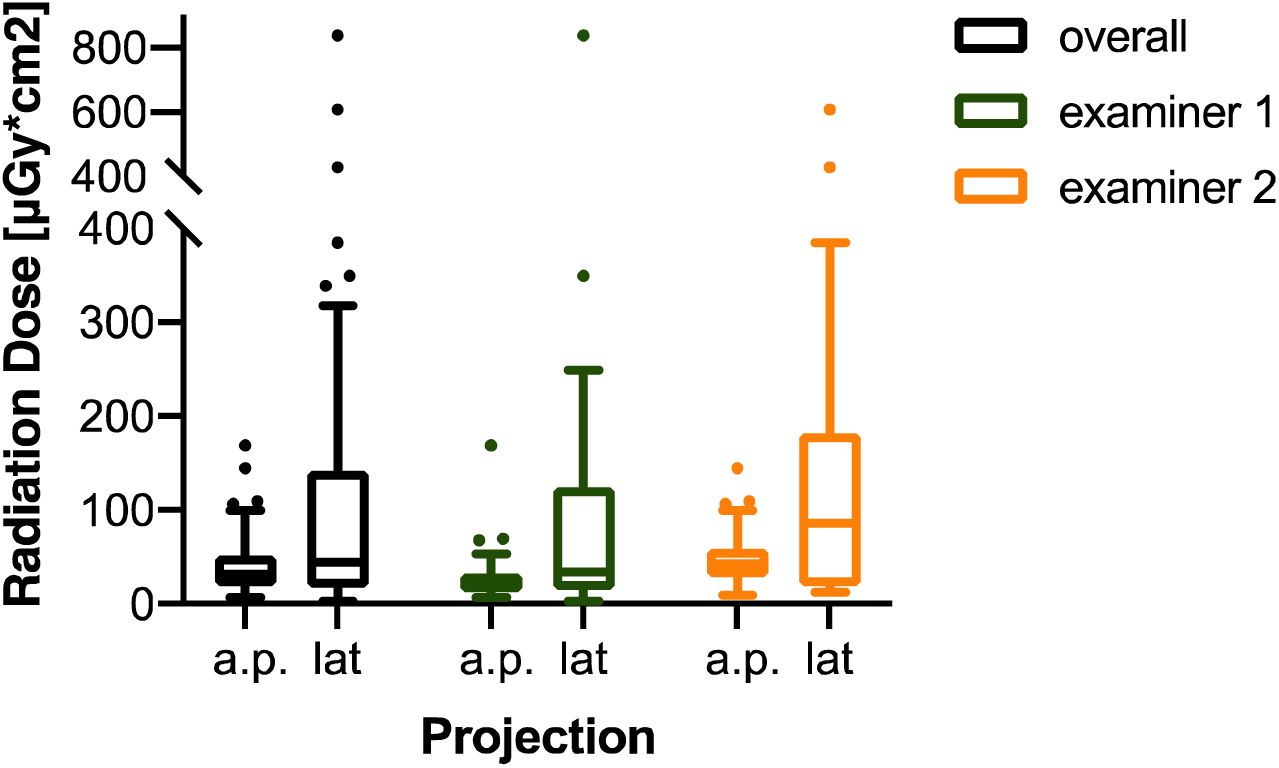
Radiation dose caused by the manual adjustment of a standard projection for a single vertebral body.

### Interrater and intrarater variance for manual adjustment of standard projections

The mean variance of the central beam θ between examiner 1 and examiner 2 was 7.6°, while the mean variance of the orbital rotation angle α was 3.8° and the mean variance of the angulation angle β was 5.4°.

For intrarater variance, the mean difference of the angles θ, α and β between the different examiners were 4.2°, 1.9° and 3.1°, respectively.

All parameters of mean interrater and intrarater variance including the 95% confidential interval are displayed in Table 2 as well as in Figure 5 and 6.

**Table 2:** Mean with standard deviation (SD) and 95% CI for interrater and intrarater variance parameters.

|  | interrater |  |  | intrarater |  |  |
| --- | --- | --- | --- | --- | --- | --- |
|  | overall | a.p. | lat | overall | a.p. | lat |
| $d\theta [^\circ]$ | $7.6 \pm 5.0$<br>(1.0-20.1) | $8.9 \pm 5.8$<br>(1.9-22.0) | $6.4 \pm 3.7$<br>(0.4-11.2) | $4.2 \pm 4.5$<br>(0.2-16.1) | $4.8 \pm 4.8$<br>(0.9-20.8) | $3.5 \pm 4.2$<br>(0.2-14.0) |
| $d\alpha [^\circ]$ | $3.8 \pm 4.1$<br>(0.1-10.6) | $3.1 \pm 3.6$<br>(0.2-13.1) | $4.6 \pm 4.5$<br>(0.0-10.4) | $1.9 \pm 2.8$<br>(0.1-10.0) | $1.7 \pm 1.1$<br>(0.2-4.3) | $2.2 \pm 3.9$<br>(0.0-10.1) |
| $d\beta [^\circ]$ | $5.4 \pm 5.1$<br>(0.5-19.7) | $7.7 \pm 6.2$<br>(0.8-22.0) | $3.1 \pm 2.1$<br>(0.1-7.1) | $3.1 \pm 4.1$<br>(0.1-13.2) | $4.1 \pm 5.0$<br>(0.1-20.8) | $2.0 \pm 2.6$<br>(0.1-9.9) |

**Fig. 5.**
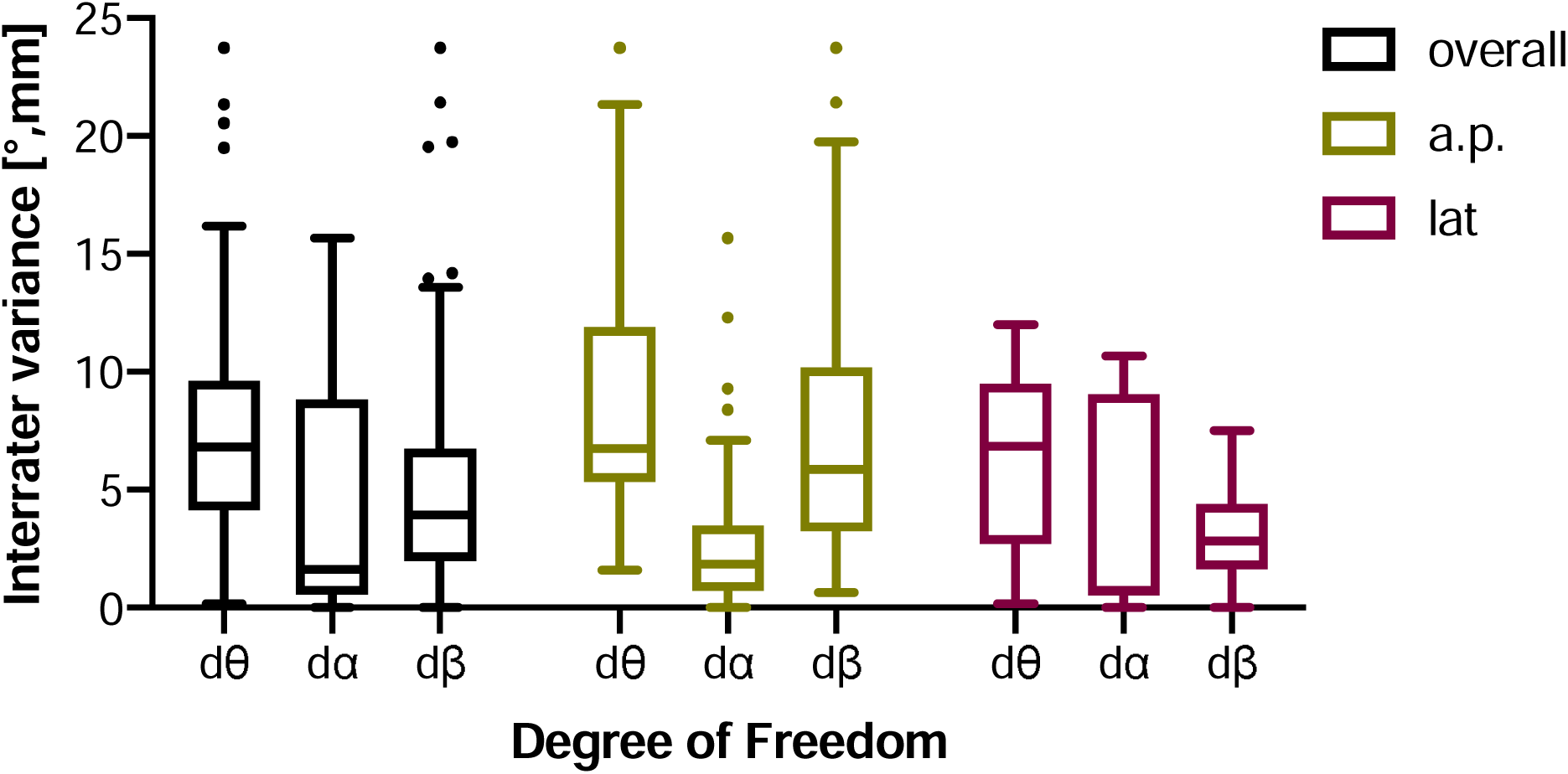
Interrater variance for the different degrees of freedom investigated.

**Fig. 6.**
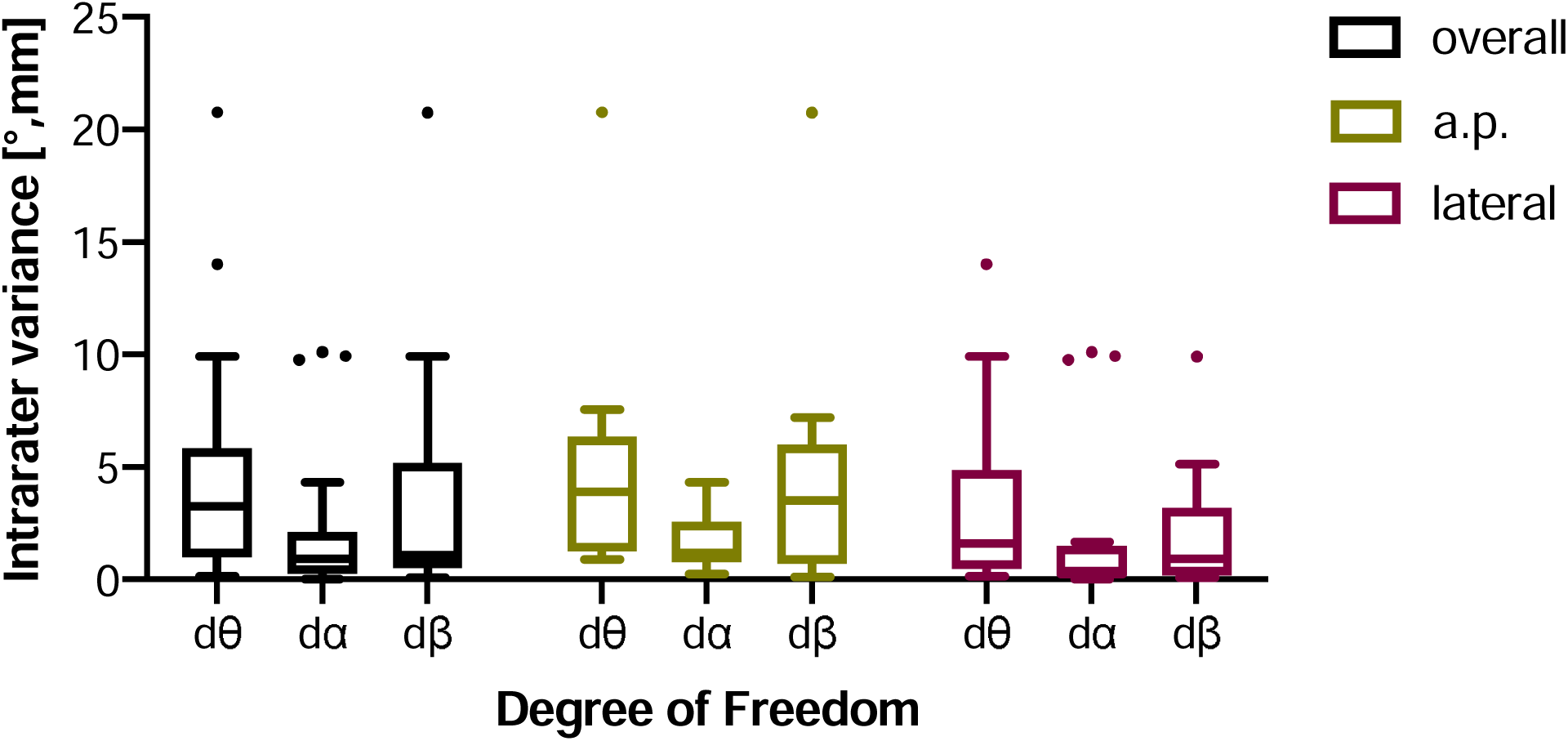
Intrarater variance for the different degrees of freedom investigated.

**Fig. 7.**
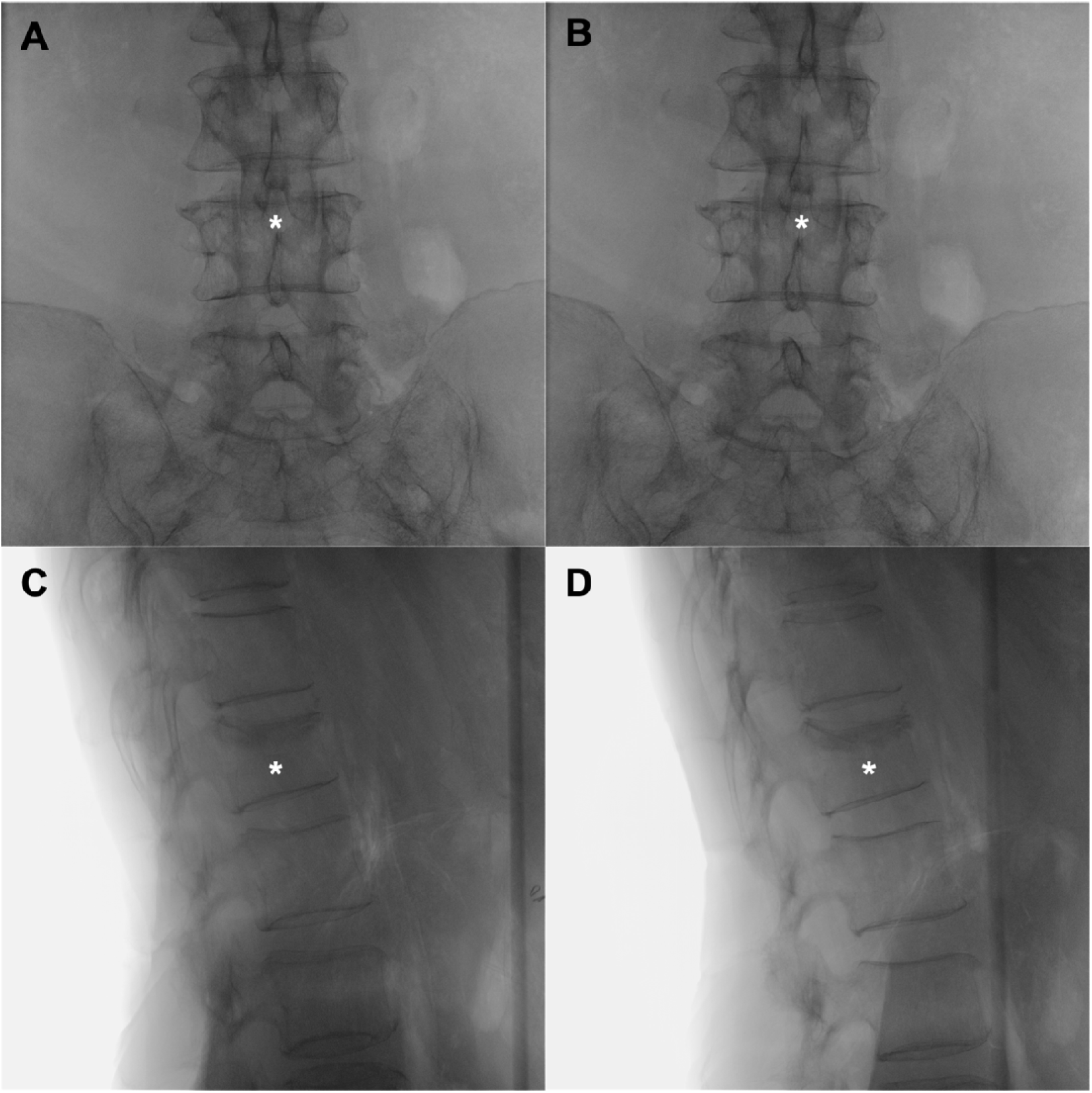
Ap standard projections of L4 and lateral standard projections of L1 as adjusted by examiner 1 (A and C) and examiner 2 (B and D). The adjusted vertebral bodies are marked with an asterisk (*).

## Discussion

The aim of this study was to assess the intra- and interrater variance for the manual adjustment of radiographic standard projections of the spine, as it is performed in operating theatres around the world every day. Additionally, we examined the amount of radiation caused during the process of manually adjusting standard projections, as well as the amount of time needed.

The time needed to adjust the standard projections was not significantly dependent on the level of experience of the examiner. There was also no significant difference for the different standard projections ap and lateral, although a tendency towards more time being required for the lateral setting was observed. Interestingly, the number of radiographs obtained was significantly higher for the ap standard than for the lateral standard. Furthermore, there was a significant difference between the two examiners, with examiner 2 requiring more images until subjective satisfaction with the projection was achieved. At this point, it is important to emphasize that the single images required for the identification of the respective vertebral body were not included in the number of X-ray images obtained. The intention was to prevent the data from being distorted to the disadvantage of the vertebral bodies in the region of the middle thoracic spine.

In summary, the experience of the examiner has only a minor effect on the time required, but it does have an effect on the number of X-ray exposures taken within this time and the resulting radiation dose. Although no measurement of the radiation exposure of the examiner was performed in the context of this study, it would be possible - in accordance with the findings of Khan et al. and Quah et al. - that the significantly higher number of X-ray exposures in only slightly more time could have played an influence on the distance between the operator and the radiation source. This would be relevant to the extent that, for a variety of reasons, radiation protection measures cannot always be implemented as consistently in the operating room as they were in this experimental study. Therefore, it is of high importance that the importance of reducing radiation exposure for both patients and staff is repeatedly brought up in the context of surgical training[8, 9].

The standard projections considered correct by the two examiners differed with regard to the central beam by an average of 7.6±5.0°, with the variance for the ap setting being slightly higher than for the lateral setting.

For the two runs performed by examiner 1 on one specimen, the intrarater variance of the central beam was reduced to 4.2±4.5°. As with the interrater variance, the variance in the ap setting was greater than for the standard lateral projection, and, again as with the comparison of the two examiners, the difference in angulation versus orbital rotation was increased for the ap projection, whereas it was the other way around for the lateral projection. These results seem clinically plausible, as these are the primary variables in the setting of each standard projection.

Comparison of our results with the literature was complicated by the limited amount of similar data.

In their study, in which different standard projections were set on the pelvis by radiology technicians and compared with a surgeon-defined ground truth, Da Silva et al. reported a variance of 2.6±2.3° in the orbital direction and 4.1±5.1° in the angulation direction. The results obtained in the present study differ only slightly from these data, although they were obtained in the spine.

Kausch et al. had two clinical experts set a reference standard for the proximal femur and the lumbar spine, resulting in an interrater variance of the central beam of up to 6.3°. For the fourth lumbar vertebra the intervariance variance was 2.5±1.6° and 1.9±1.3° for ap and lateral projections, respectively. Considering that this reference standard was set by adjusting planes in a 3D data set and subsequent comparison of the central beam angle, our results from a simulated clinical scenario seem reasonable[7]. In further experiments, Kausch et al. determined a mean interrater deviation of dθ = 6.1±4.2° for the manual adjustment of standard projections for the forth lumbar vertebra. Using their proposed approach for C-arm pose estimation with only one single image needed, they outperformed the manual adjustment on all six specimens used[10].

In the aforementioned study by Da Silva et al. it is stated that an average of 8.0±4.5 exposures were required before the results were accepted by the investigators[2]. In a different study performed by the same authors, an average of 6.4 radiographs was necessary to adjust the standard projections in an experimental setting. This value is confirmed by the retrospective analysis of clinical data of the same region with an average of 6.6 ± 4.9 (range 2-14). It was not possible to determine whether the data on which the two publications are based overlap.

The authors state an average value of 41.0±30.9 seconds as the time required for positioning the C-arm, although information on the exact collection of these values is missing, which makes comparison with our study difficult[1].

Nevertheless, the comparison with the literature shows that the results we presented from the spine can apparently be transferred to other regions with complex anatomy, which increases the significance of our data as well as the clinical relevance of the subject discussed.

The validity of the study is limited by the fact that only two of the six degrees of freedom of the C-arm are examined in this study. However, these are the two variables that are most relevant for the adjustment of standard projections of the spine. The investigation of the interrater variance with regard to the swivel was omitted, since this is of only secondary importance for the setting of the standard projections in this anatomical region and was accordingly not changed in the course of the experiments.

It would certainly have been desirable to investigate the intrarater variance with respect to the different experience level as well, but this was not possible due to the limited time that the specimens were available for investigation.

Another limiting factor to mention is that the classification of the results is hampered by the limited availability of similar studies in the literature. Apart from the discussed results of Kausch et al., which were also performed in cooperation with our research group, there are no data on interrater and intrarater variance for the manual adjustment of standard projections in the spine. For other anatomical regions as well, the number of publications is extremely limited. This also applies to data regarding the consequences of this standard process performed in hospitals around the world every day. In this regard, we are also the first to quantify the number of radiographs and the time required for the anatomical region of the spine.

## Conclusion

In summary, this study investigated the interrater and intrarater variance for standard manual level setting in the thoracic and lumbar spine. Additionally, we were able to quantify the time and number of radiographs required for this procedure for different levels of experience, as well as the resulting radiation dose.

## Data Availability

All data and statistics are available on reasonable request from the corresponding author.

## Acknowledgements

The authors would like to thank Rimasys GmbH for providing the human specimens as well as the facilities for conducting the experiments.

